# A Dense sub-graph based approach for Automatic detection of Optic Disc

**DOI:** 10.1101/2022.06.27.22276966

**Authors:** Subrata Jana, Tribeni Prasad Banerjee, Gour Sundar Mitra Thakur, Pabitra Mitra

## Abstract

Glaucoma is a situation of greater than normal intraocular pressure surrounded by the eyes. This explains the harm to the optic nerves as the limb passes in sequence to the brain. The graph base is used in this paper for automatic localization of the optic disc. This paper proposed and modified a new dense sub - graph approach to locate the affected optic disc by using DRIVE, STAIR, and Drishti -GS1 databases. This model has proved to be more accurate when compared with other standard models representing the concert’s progress. This method provides a new idea for the location of the optic disc with a system accuracy of 93%.

## 1 Introduction

Glaucoma infection is a general infection in ophthalmology. This is primary optical infection. Many infections will have different symptoms on the fundus, such as diabetes, hypertension, and so on. The main physical structures are the fundus image, the group of blood vessels, the optic disc, the optic lens, and the mark. The position of the optic disc is helpful to the consequent position of the mark. The optical disc location is primarily based on the following characteristics of the optical disc: One of the features of optic discs is their round or oval shape. Another feature is that the retina blood vessels are either thick or thin from source to end. The blood vessels on the entire retina may be roughly parabola shaped. Blood vessel circulation in the optic disc area is vertical, whereas blood vessel circulation in other areas is horizontal. People are expected to perceive visually idiosyncratic, or salient, scene areas fluently and quickly. This filter area is then purged and the procedure is better understood for the removal of the comfortable high level in sequence. The ability has extensively been considered by cognitive scientists and has recently fascinated a set of awareness in the computer vision society, mostly because it assists in locating the object [1] or area that proficiently a view and thus controls multifarious vision troubles such as view accepting. The goal of this research is to locate the optic disc [2, 3] in order to remove the prominent area in an image by combining excellent pixel segmentation and graph theory based saliency area detection. Dense K-sub-graph constructions are oppressed to acquire and improve saliency diagrams. First, we screen the image into the areas using the SLIC [4] segmentation method. The graph-based Markov chain [5] random walk[6] model using saliency model features is color, compactness, and intensity. The SLIC method generates a different sparser graph on which the k-dense [7] sub-graph is worked out. The clustering algorithm is beautiful for the assignment of class detection. The application to a large spatial database increases the clustering algorithm’s subsequent constraint.

1. Sphere data to decide the input constraint since suitable values are repeatedly not identified in progress when selling the big database.
2. The detection of the cluster with a random contour is necessary because the contour of the cluster in a spatial database may be circular, long-lasting, linear etc.

We present the SPDBSCAN [8] clustering algorithm. It requires four input parameters and supports the user in determining an appropriate value for them. It determines the cluster of dissimilar contours. SPDBSCAN is a well-organized level for big databases. A number of graph-based saliency algorithms are well-known in the literature to help improve on the contrast determined to represent the inter-nodal evolution probability [9], present superior random walk measures like random walk, or use various entropy functions [10]. Most graph-based techniques construct a fuzzy saliency map. The most salient region [11, 32] is useful for saliency mapping to filter. Our model is a graph-based technique that uses dense sub-graph calculation and sorts out the most important areas after a random walk. Salient regions and non-salient regions are combined; the deferred saliency map gives better results than any other existing method. This has been realised by taking into consideration an extra informative local graph construction, i.e., a dense sub-graph, rather than easy centrality evaluation in finding the map. A dense k-sub-graph based algorithm for compact salient image region detection [12, 32] using the saliency model. This technique does not provide better results for optic disc location, but we have modified the slic technique and use spdbscan to get better results than only the saliency technique. The rest of the paper follows the order: Section 2 describes related work; Section 3 Proposed Methodology, Section 4 Graph based Saliency Detection, Section 5, Describes experimental results; Section 6, tests plot graphically; and Section 7 conclusion and future work.

## 2 Related Work

Saliency computation is a deep-rooted psychosomatic hypothesis about human concentration, such as Feature Integration Theory. The hypothesis confirms that a variety of facial expressions are method in equivalent in unusual areas of the human brain, and the position of the facial appearance is combined in master map position and concentration selects the existing region of concern. It is used for arbitrary shape of clustering using DBSCAN [8]. It is a density-based technique. This algorithm gives better results and more efficiency. Graph Based Visual Saliency Model [9] detects high salient regions and upper saliency values in the image plane. The dissimilar methods used to develop the unprocessed fundus images, such as the difference improvement, clarification rectification, mask production, and the technique used for segmenting the optic disc, were categorized into possessions based vessels junction and pattern matching methods. As Site Entropy Rate (SER) [10] supports the rule in sequence maximization, saliency finding in both still and video images, visual saliency is definite. This model estimates the conversion probability among two graph nodes by fusing the difference in sub-band facial appearance answers and the spatial distance with a pre-set limitation. The graph based saliency discovery technique is relevant on segmented images established by the SLIC super pixel segmentation algorithm and k-dense [12] sub-graph ruling problem to that of saliency detection to get a better drawing out of salient parts in the illustration in sequence. We can find salient objects [15], which is prepared as an image segmentation using a set of local, area, and global salient objects. This finding has wider uses. Physically assembling and preparing images for object detection is very costly. The bottom up saliency decision method with incorrect border [16] line elimination and normalized random walk ranking and incorrect border line deletion process successfully abolishes the image border line, with the border line adjacent to the centre super pixels. It is a bottom-up [17] loom using a low level feature. It is also a multi-state loom where preferred scales are needed. The visual saliency finds nodes in the most important demonstration, and quantifiable representation of hierarchical [18] entity based concentration for computer visualization. It is entity-based and spaced-based concentration can be incorporated by using assemblage-based salience to covenant with lively diagram task. The task saliency based area selection gets better object detection in high chaotic [19] scene reflection and to guess human fixation on images, and confirms the pressure of numerous scales, color space weights, and quaternion change axes. The global and local possessions of a region can be explored by hauling out random walks on an entire graph and a k-regular graph [21].The cellular automata [22] technique to create a conditions-based map, which obtains both global color and spatial space matrix into contemplation based cellular automata instinctive renew method, is planned to utilize the fundamental connectivity of salient objects throughout communications with neighbors.. The GBVS provoked [9,23] centre bias by making active then standardizing an identical image using this algorithm. They also discuss how the typical algorithm guesses fascination better but still poorer than GBVS. The visual consideration necessitates three different stages. Firstly, a set of basic features is calculated in equivalent crossways the illustration field and is symbolized in a set of cortical topographic maps. These maps are united into the saliency map, training the relative conspicuity of the visual scene. Next, the WTA system working on this map singles out the most prominent location. Next, the assets of the select position are routed to the essential illustration. The WTA network then moves robotically to the most prominent place to show the facility to successfully shrink [25] the alter times and hunt errors to choose helpful things, regions, and prepared groups, and the capability to elastically choose visual things whether positioned in the fovea field or on the diagram fringe. The finding of the optic disc [26] location and function of the macula also discusses corner detection, the optic disc location convoluted, the corner detection complex and the computational tool for automatic [27] glaucoma detection and thresholding measure cup and segmentation ratio. The dense dilated element removal block into an encoder-decoder [28] configuration to remove accrued attributes at different scales and an optic disc and optic cup segmentation technique using the Graph Convolution Network (GCN) [29] method. G_Net, C_Net proposed two new techniques to locate the optic disc and optic cup, and iris segmentation using IsqEUNet [34]. This technique has good results. Other than the PCA and Hessian Method, vessel segmentation using the max-flow graph [33] yields better results.

## 3 Proposed Methodology

The block diagram of the optic disc location is exposed in Fig. 1. The structure has been separated into three steps, that is to say, The three stages are: Feature Extraction stage, Training Stage, Testing and Evaluation Stage.

**Fig 1.**
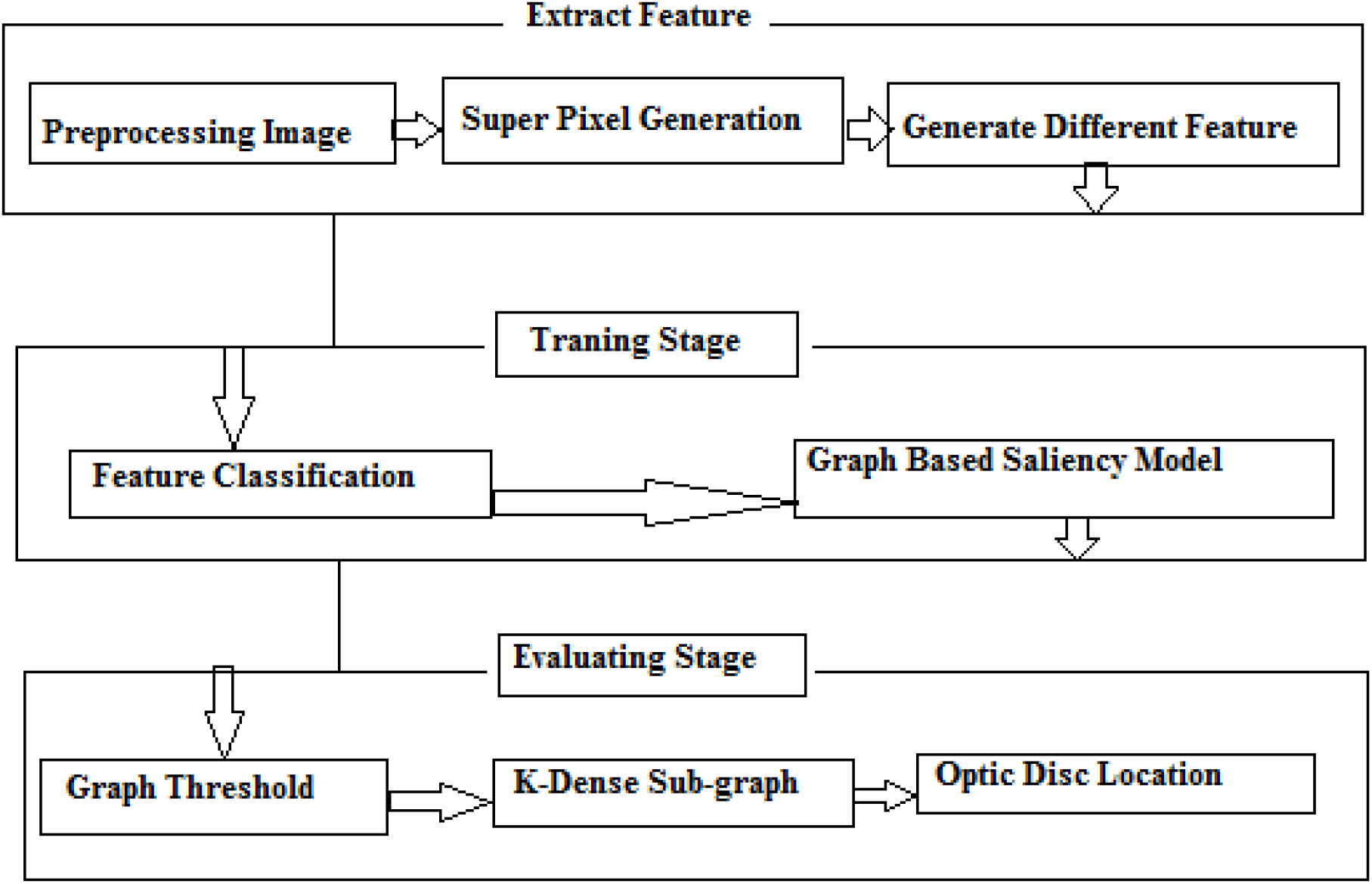
Block Diagram of Optic Disc Location Framework

Step1: The first stage is the image region or super-pixel, generated from the original image using the SLIC technique.

Step2: We use an extract feature map graph based saliency model and a compactness factor for saliency map computation.

Step3: We apply a graph based edge threshold in sparse graph construction.

Step4: The last stage is the evaluation stage. At this stage, we apply k-dense subgraph computation, which results in a salient region.

It may be practical from Fig. 2 that we remove the element in sequence and then use the graph based saliency model to get an in-between saliency map, which is extra polished by dense sub-graph calculation, to attain the ultimate saliency map. This technique creates the connectivity graph based on super-pixel, which calculates the graph based on a rectangular region. The HSV color space was chosen because the Euclidean distance in this color breathing space is uniform, and it has been experimentally proven to produce better results than YCbCr and RGB.

**Fig 2.**
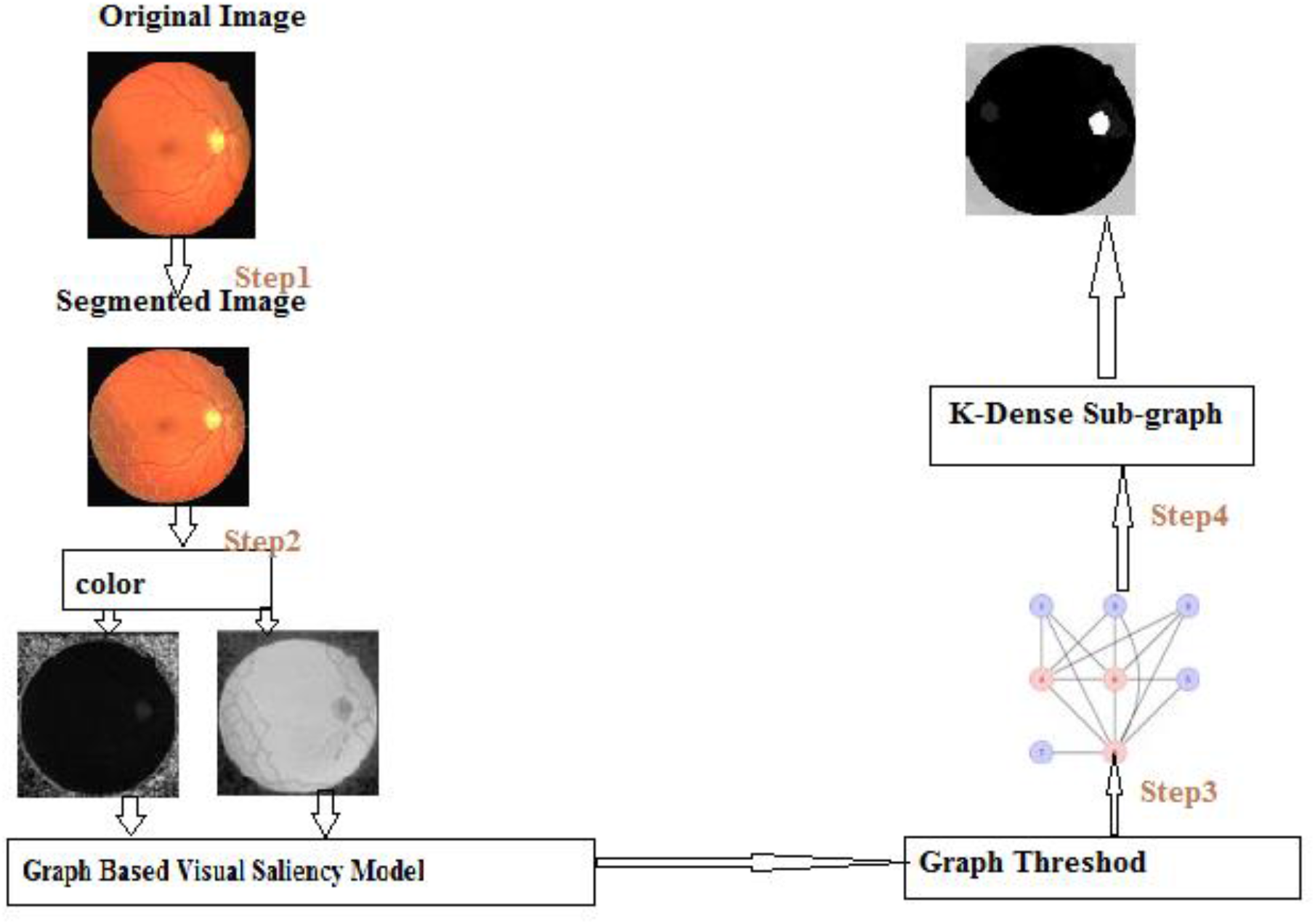
Flow Chart for Modified Dense Sub-graph technique

**Fig 3.**
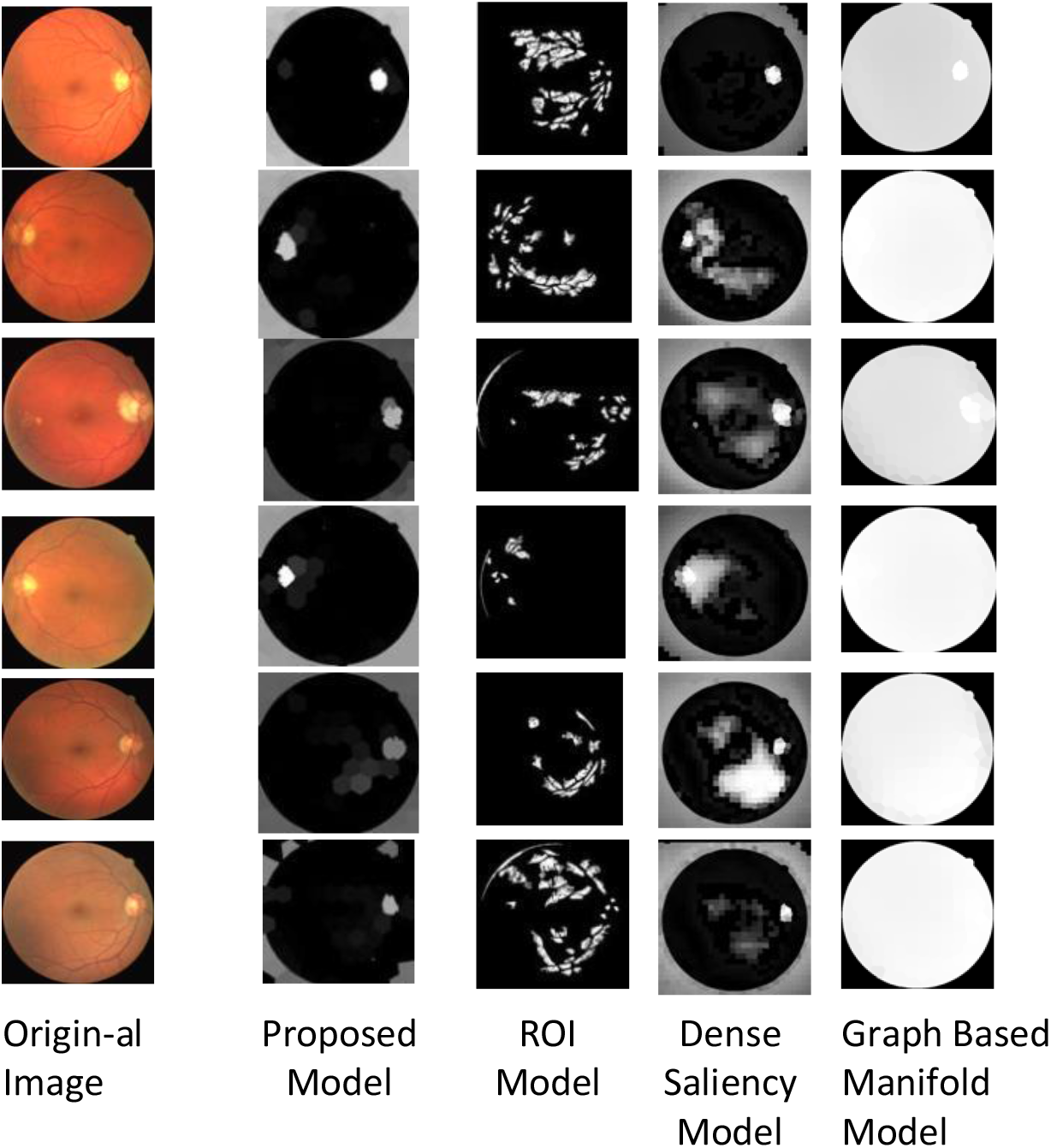
Comparison of saliency map different model

**Fig 4.**
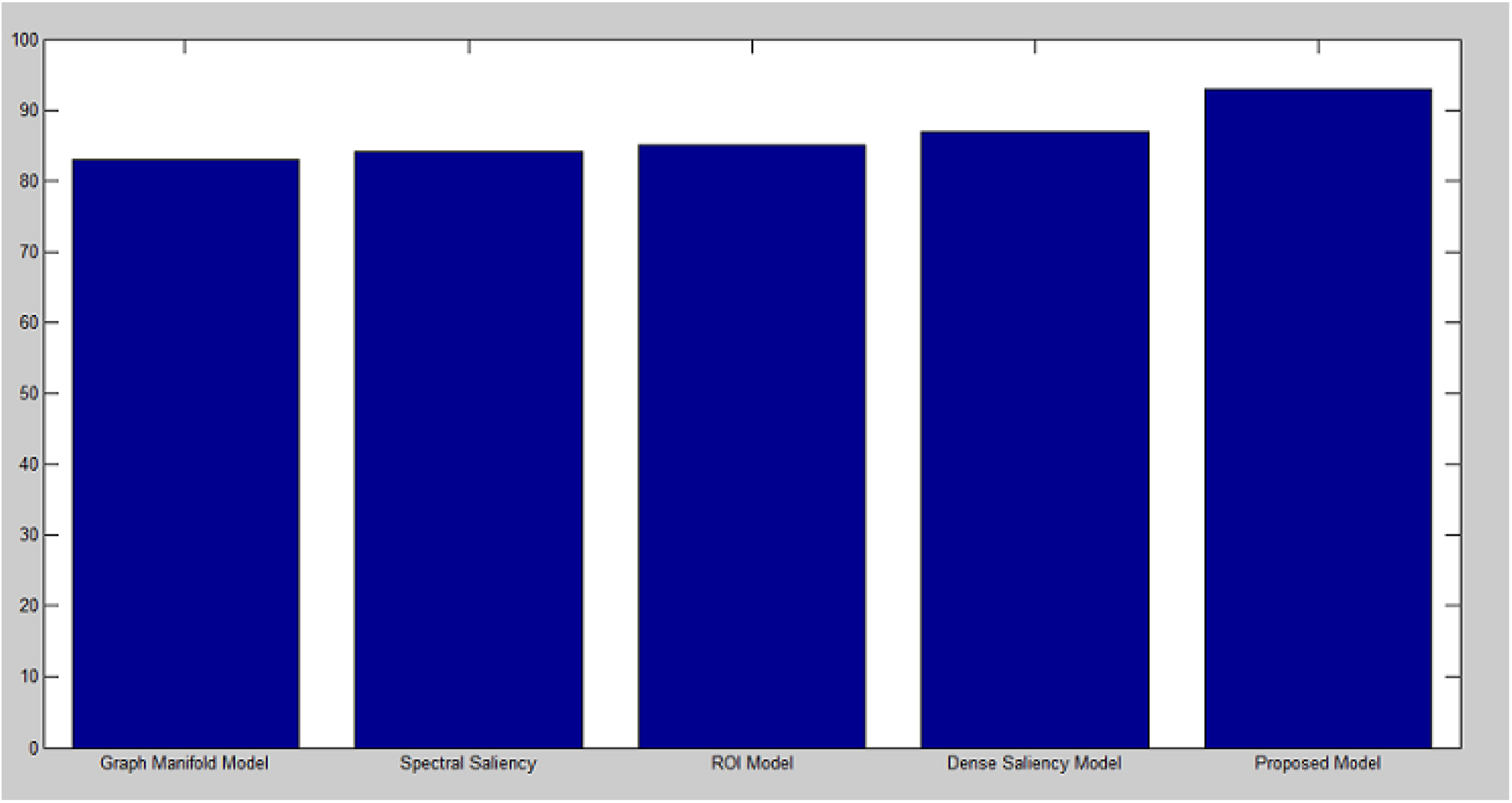
Graphically represent different techniques

### 3.1. Algorithm

SLIC stands for Simple Linear Iterative Clustering, it is used our modified SLIC[4] technique. This algorithm accepts 6 parameters. First parameter is input image denoted *im*, second parameter is *k* that is preferred number of super pixel same size, we consider say super-pixel 2000, third parameter is *m* that is use smoothing shape, fourth parameter is *seRadius* that is denoted adjacent region, fifth parameter is *colopt* indicating how the color center to be computed, sixth parameter is *mw* indicating the window size, it is optional. We can used initial cluster center D_i_=[l_i_,a_i_,b_i_,x_i_,y_i_]^T^ and usual grid space denoted S. We construct approximately same size super-pixel, space gap is 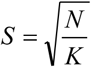. The Cluster center are stimulated the minimum gradient position in a 3×3 quarter. We use this technique reduce noise of super-pixel. K-means clustering each pixel compared to all clustering center where as super-pixel clustering each pixel compared small region super-pixel center. Super-pixel is an area approx SxS, correspondingly super-pixel center region is approx 2S x2S and measuring Euclidian distance denoted D. All the pixel belonging to the nearest cluster center and update cluster center D_i_=[l_i_,a_i_,b_i_,x_i_,y_i_]^T^ and usual grid space denoted S. The Cluster center are stimulated the minimum gradient position in a 3*3 quarter.

#### Algorithm

Initial Cluster center D_i_=[l_i_,a_i_,b_i_,x_i_,y_i_]^T^ and grid space denoted S

Lowest 3×3 neighbors position grid space.

Step1: Set label(i)=-1 Set distance p(i)=*α*

Repeat

Step2:While cluster center D_i_ do

Step3:While pixel j in 2S*2S and cluster center C_k_ do

Step4:Calculate Euclidian distance P between D_i_ and j

If P<p(i) then

P(i)=P

L(i)=i

End if

Step5:Calculate new cluster center

Until *ξ* <=thresh.

The SPDBSCAN function determines the neighborhood of any super-pixel. The following criteria are used: Any two super-pixels are adjacent. The clustering distance measure is the lab color distance between the color centers of two super-pixels. If any two super-pixels are not adjacent, the clustering distance is measured to be infinite.

SPDBSCAN required four parameters: lm, Sp, Am, and E. The first parameter, lm, denotes the label image generated by the SLIC function. The second parameter, Sp, in each column gives the attributes of each super-pixel region generated by SLIC. The third parameter, Am, is the adjacency matrix of the label image. The fourth parameter is E, which denotes the threshold that controls which super-pixels are clustered together. In our program, we consider the threshold value E to be 5. SPDBSCAN function to find the cluster beginning with an arbitrary starting point Sp and rescue all points. If m1 is a boundary point, do not consider this point. SPDBSCAN can take the next point. SPDBSCAN used two global variables. SPDBSCAN combines two clusters into a single cluster. Let the distance between two sets of points M1 and M2 be what we can describe as *dist*(*M*_1_, *M*_2_) = min {*dist*(*p,q*) | *pεM*_1_, *qεM* 2 }

Input: Initial this function takes label image of super-pixel, Adjacency matrix of segment, structure array of super-pixel

Output: New cluster region

Step1:Set Np=length(structure array of super pixel)

Step2:While n<Np do

If n<> visit

Visit(n)=1

Step3:Neighbours=regionquery(Sp,Am,n,Ec)

Step4:While ind<=Neighbours do

Step5:Nb=Neighbours(ind)

Step6:neighboursP = regionQueryM(Sp, Am, nb, Ec);

Step7:neighbours = [neighbours neighboursP];

If the super-pixel has not been visited yet, mark it as visited, then find its neighbour and increase the number of clusters. If the neighbour has not visited, mark it as visited and add it to the list. If the neighbour is not a member of the cluster, add them to the cluster list.

### 3.2 Super-pixel Segmentation and Feature Extraction

The first image is segmented using the SLIC method, takes 6 input parameters such as *im* for input image and *k* for the number of desired super-pixel. This is nominal; the actual number of super-pixel generated will generally be a bit larger. In our program, *k* value of 100 pixels per super-pixel is used. The next parameter is m, which is the weighting factor between color and super-pixel. We can now define the value of 25.parameter is *SeRadius*. Regions morphologically smaller than this are merged with adjacent regions. *The SeRadius* range is 1 to 1.5. In our program, we can define value as 1. The next parameter is *colopt*, indicating how the cluster colour centre should be computed. In our programme we can use the median. The next parameter is mw. It is an optional median filter window size. The slic method returns l, Am, and c, where l is a labelled image of a super-pixel, Am is a segment adjacency matrix, and c returns a super-pixel attribute structure array with fields L, a, b, r, c, stdL, stda, stdb, N, edges, and D. This step is SPDBSCAN, or the Super Pixel Density Base Spatial Clustering Application With Noise method. This function is used for clustering for image segmentation. This function takes four parameters: *l, Cp, Am, and E. Now to explain the* first parameter *l*, labelled image of clusters/regions generated by a slic method. The second parameter is *Cp*. Each column gives the attributes of each super-pixel region. Its value reruns the slic method. The next parameter is *Am*, an adjacency matrix of the labelled image. Its value is returned by the slic method. Value is *E*, the matching tolerance value/distance threshold that controls which super-pixels are clustered together. In our program, we defined value 5. This function returns The following function is DRAWREGIONBOUNDARIES, which displays the drawn boundaries of labelled regions in an image. Channel denotes a hue image, the S channel denotes a saturated image, and the V channel denotes a value (intensity) image. We only use the Hand S channel.

## 4 Graph Based Saliency Computation

Image segmentation is accomplished through the use of the SLIC, SPDBSCAN, and DRAWREGIONBOUNDARY approaches. Now we’ll make a graph. Image by taking into consideration segmented image area as vertex and aloofness. Graph Base Visual Segmentation united the loads as the product of different loads [12].

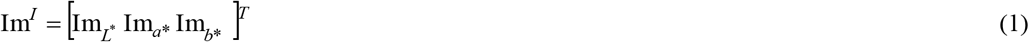

Where Im_L*_,Im_a*_,Im_b*_ are the regularized attribute strength plan matching to L*,a*,b* apparatus of the image and Im^I^ is a vector including three features map.

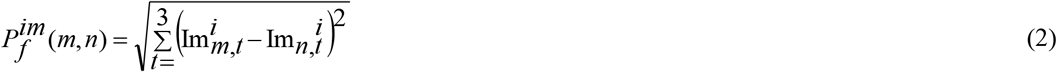

Where 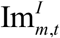 and 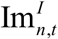 are mean intensity assessment of the element canal t,where t=1,2,3 for L*,a*,b* consider for nodes m,n correspondingl [12].

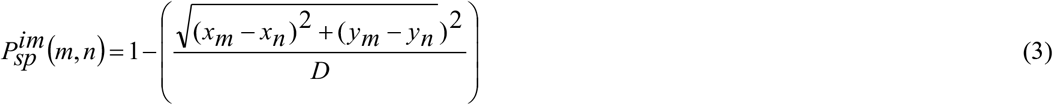

Where x_n_ and y_n_ symbolize the mean x and y co-ordinates and D represent diagonal length of the image. M_GBVS_ is a fully connected graph, whereas G_GBVS_ is based on the saliency map. Now we calculate the K dense sub-graph of this graph. The graph G_im_ derived from N*N transition matrix, where N is the number of nodes. The element TP(i, j) is proportional to the graph weight w(i, j).The graph G_image_ is a diagonal matrix whose degree matrix which is denoted W,the equation is[12]

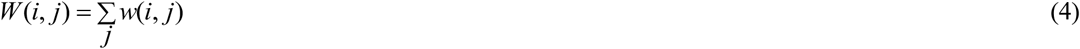

Each column sum of the value in transition matrix must be 1.the transition matrix equation is [12]

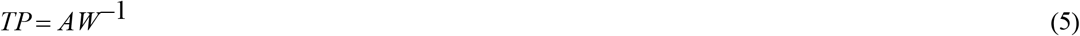

A random walk from the Markov chain is used [22]. G_image_ has a fixed number of nodes and is fully related by construction to the stationary distribution for Markov chain exit. M_GBVS_ is an entirely connected graph using the G_GBVS_ saliency graph. The K-dense[7] subgraphs calculate the solidity of the graph, using the threshold edge.

### 4.2 Saliency Graph Thresholding

The saliency map shows different feature values. Dissimilar features are proportional to edge weights. In saliency computation, they require higher edge weights. A threshold partitions the graph where more thresholds accept the graph and fewer thresholds discard the graph. The distribution of edge weights between all pairs of vertices is analyzed using threshold T. We can create two sets of edges: one set is the reject set, denoted Pr, and the accepted set is Pa. Distribution detect threshold has been used in entropy. For a particular threshold t, the ratio r of the summation of weights for the rejected set Pr of edges to the total set Pr U Pa, of edges is calculated as [12]

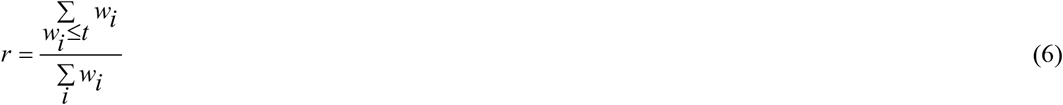

Above Equation (6) returns the selected set of edges to the total set of edges. En denotes the rejected and selected set of edges. The equation given below [12]

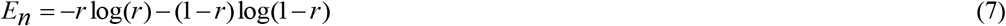

The threshold t varies with the edge weight entropy rate. En. The edge weight threshold T_max_ is chosen, then the edge weight entropy maximum. My experiment threshold period is.1, where the starting threshold Ti =.1 and the end threshold Tf =.95. The highest max-entropy value is.69, the maximum threshold value is.44, the ration rejection value is.03, and the ratio accepted value is.97.

### 4.3 Dense k-Subgraph Computation

A graph is a collection of vertex and edge groups in which the densest k-subgraph rules a sub-graph precisely k vertices, i.e., the maximum number of edges. It is also known as the NP hard problem. The dense k sub-graph problem can be formulated as given below [8].

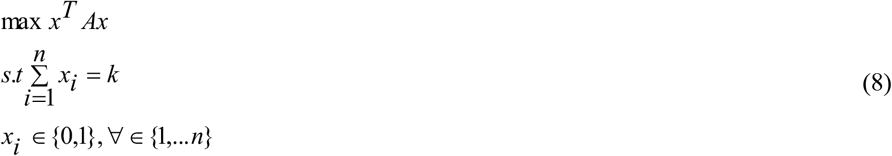

In the above equation, A is the adjacency matrix and k is a positive integer between 3 and n-2, and xi is a binary variable that obtains value one.

Let f(x)=x^T^Ax and n=N with each step coordinate value renewing. In each step I,let Ti,|Ti|=m the set of random coordinate renew concurrently. Each iteration, the random coordinate value is updated following the equation [8]

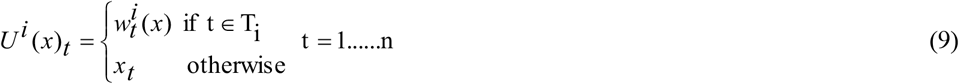

Where *w*^*i*^ (*x*) ∈*ϕ*^*m*^ solution is dished optimization dilemma.

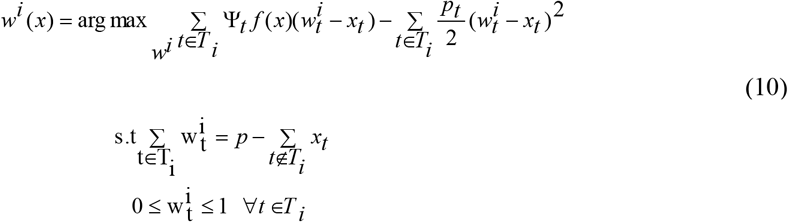

In above equation (10) solved linear programming problem.

### 4.3 Saliency Map Computation

The map follows the sequence of steps.

Step 1: Let Ssalience = v1, v2, v3,… vn represent the vertices’ positions in a subgraph, and Msaliency (x, y) represent the saliency value at a pixel M(x, y), where x and y are pixel positions.

Step 2: In step 2, the pixel value corresponds to vertex I for the image’s pixel. If I establish that the mean vertex degree is compared with the degree of vertex I and assigns value in saliency. If I do not establish Msaliency (x, y), its value is zero.

Step 3: For the final step of the saliency map, we use the equation [11].

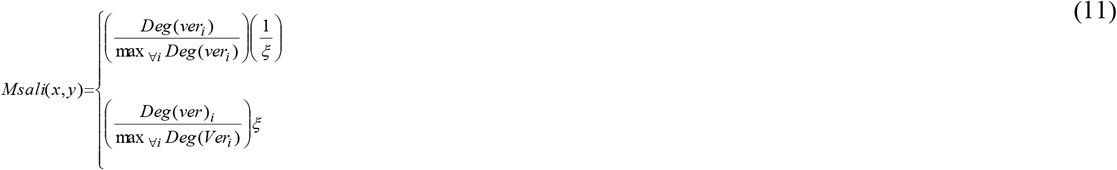

In the above equation *ξ* < 1,the pixel section of the saliency value has an association to a low degree. On the other hand pixel communicates with a high degree of allocation of greater saliency value.

## 5 Experimental Results

We used the most popular datasets, such as the DRIVE and STAIR datasets. We have implemented our algorithm and other models. In our algorithm, two parameters are required: one k-dense k, and where k-dense subgraph determines the region of saliency value, which is an improvement part and maps to the correct superiority. We contrast our model with other four models such as the Graph Based Manifold Model [13], Dense Saliency Model [12], and ROI model [14]. We can see that our model outperformed the other four models. We can use a large number of super-pixel nodes (n = 100).

We use a large value to implement a super-pixel smoother shape. The m value is 25, and we use the seRadius value is 1. We experimental that k=.8%n better result than other method. We discovered that *ξ* ≺ 1 produced a more visible result than other existing techniques. We choose *ξ* value of.3 for prominent results.

## 6. Text Plot Graphically

Accuracy concludes that how proportion of test data is properly secret. It can be calculated according below this equation

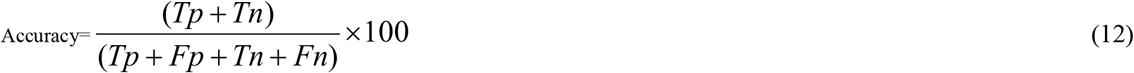

**TABLE I.** ACCURACY RESULT

|  | Model Name |  |  |  |  |
| --- | --- | --- | --- | --- | --- |
|  | *Graph Manifold Model* | *Spectral Saliency* | *ROI Model* | *Dense Saliency Model* | *Proposed Model* |
| Accuracy(%) | 83 | 84 | 85 | 87 | 93 |

Tp denoted True Positive,Tn denoted True Negative,Fp denoted False positive value, Fn denoted False Negative value.

Graph Manifold model[13] avg accuracy 83%,Spectral Saliecy multichannel model[31] avg, accuracy 84%, ROI model[30] avg. accuracy 85%. dense saliency Model[12] avg. accuracy 87% and our model avg. accuracy 93%.

## 6. Conclusion and Future Scope

We have proposed a new technique for salient region detection. Our primary goal is simple linear iterative clustering, followed by image segmentation via SPDBSCAN from the super pixel cluster. It is more efficient to determine the neighbourhood of each super pixel. It is faster than other exciting techniques. It gives better results than other techniques. The experimental results show that regulating two parameters differently than our method produces prominent results. Future work includes convolution neural network detection and noisy image datasets.

## Data Availability

All data produced in the present study are available upon reasonable request to the authors

